# FKBP51 levels negatively correlate with Depressive symptoms in Chronic Hepatitis C

**DOI:** 10.1101/2020.03.19.20039248

**Authors:** Anh Duy Pham, Darren Wong, Sern Wei Yeoh, Diana Lewis, Ting Ting Lee, Md Shaki Mostaid, Gursharan Chana, Chad Bousman, Alex Holmes, Michael M Saling, Danny Liew, Alexandra Gorelik, Ian Everall, Siddharth Sood, Amanda J Nicoll

## Abstract

**Background and aims:** Patients with chronic hepatitis C infection have high rates of major depressive disorder. The reasons for this are multifactorial, but social and demographic factors do not entirely explain the increased burden. Direct neuropathologic effect of the virus in the development of depression has been postulated but the mechanisms remain unclear. Single nucleotide polymorphisms (SNPs) in FKBP5 (protein product FKBP51), a co-chaperone of the glucocorticosteroid receptor, have been associated with greater severity of affective disorders. We examined the interaction between FKBP5 SNPs and chronic hepatitis C infection in patients with and without depressive symptoms.

**Methods:** forty-one subjects completed quality of life and psychiatric questionnaires. Thirteen patients were classified as depressed on the Hospital Anxiety and Depression Scale depression sub-score (HADS-D ≥11). FKBP51 protein expression, FKBP5 mRNA and FKBP5 SNP analysis was compared between those with and without depression.

**Results:** There were no statistically significant differences between the groups in demographics, medical co-morbidities or substance use. A moderate negative correlation (Spearman rho -0.53, p<0.001) was found between HADS-D sub-score and FKBP51 protein levels in serum. Correspondingly, the average expression fold change in peripheral blood FKBP5 mRNA relative to a reference gene was lower in the depressed group at 0.76 compared to controls at 1.40. There was no differential expression of FKBP5 SNPs between the groups.

**Conclusion:** Levels of FKBP5 mRNA and FKBP51 are lower in hepatitis C patients with depression and further exploration of this interaction is required.

## INTRODUCTION

The prevalence of depression amongst patients with chronic hepatitis C (CHC) is high, with Australian data suggesting that 27 to 53% of patients with CHC have a depressive disorder compared to the lifetime prevalence of 11.6% in the general population.^1,2^ The mental health burden among patients with CHC appears to be independent of confounding factors such as substance abuse^3^. This posed a major barrier to treatment in the era of pegylated interferon. Whilst the introduction of direct-acting antivirals has mitigated much of this obstacle due to shorter duration of therapy and better side-effect profile, depressive illness may still pose a barrier to engagement with health care services and thus reduce access to new therapies^4^.

Complex biologic and environmental factors interact in the pathogenesis of depression in patients with CHC, but there is accumulating literature demonstrating the machinery exists for the virus to exert a direct neuropathic effect. Firstly, hepatitis C RNA has been isolated in the cerebrospinal fluid of those with chronic infection^5,6^. Secondly, the endothelial cells of the blood-brain barrier express the necessary HCV receptors for viral ingress^7^. Indeed, hepatitis C RNA has been isolated from brain tissue (cerebellum, medulla, white and grey matter). Additionally, cerebral metabolism is altered by CHC infection, with studies employing magnetic resonance spectroscopy demonstrating significantly higher choline/creatine ratios in the white matter and basal ganglia as well as and high levels of myo-Inositol in infected patients^8^. These changes reverse following successful clearance of hepatitis C^9^. How the virus might directly induce depression once it has entered the central nervous system is not fully understood. One possibility is through interaction with the hypothalamic-pituitary-adrenal axis (HPAA). Inappropriate, prolonged hypercortisolaemic stress response is a recognised component of biologic depression and antidepressant pharmacotherapy has been shown to normalise cortisol levels^10^.

A large body of research has been devoted to identifying the biologic underpinnings of HPAA dysregulation in depressive illnesses. Gene wide association studies have identified FKBP5 (also known as FK506 binding protein 5) on chromosome 6 as one of several candidate genes for the development of depression^10^. It encodes for FKBP51, a 51kDa member of the immunophilin superfamily that acts as a co-chaperone of a large multiprotein complex that regulates the activity of the cytosolic glucocorticosteroid receptor (GR) (figure. 1). The chaperone/co-chaperone composition of this complex alters the sensitivity of the glucocorticoid receptor to cortisol. Under physiologic stress conditions, the neuronal cytoplasmic glucocorticoid receptor translocates into the nucleus in response to circulating cortisol and upregulates gene expression to cope with physiologic stress. The FKBP5 gene itself is one such target of the activated glucocorticoid receptor. When transcription is upregulated and FKBP51 product is present on the multiprotein-GR complex, glucocorticoid receptor sensitivity is reduced^11^. FKBP5 is therefore one of the GR-dependent ultrafast negative feedback mechanisms that feedback on the HPAA to counter the cortisol response^12^. When there is excess of FKBP51 however, such as demonstrated with certain ‘high induction’ FKBP5 polymorphisms, greater FKBP51-mediated inhibition of the GR occurs and the normal negative feedback processes are not upregulated; thus a maladapative hypercortisolaemic response is induced^13^.

**Fig. 1.**
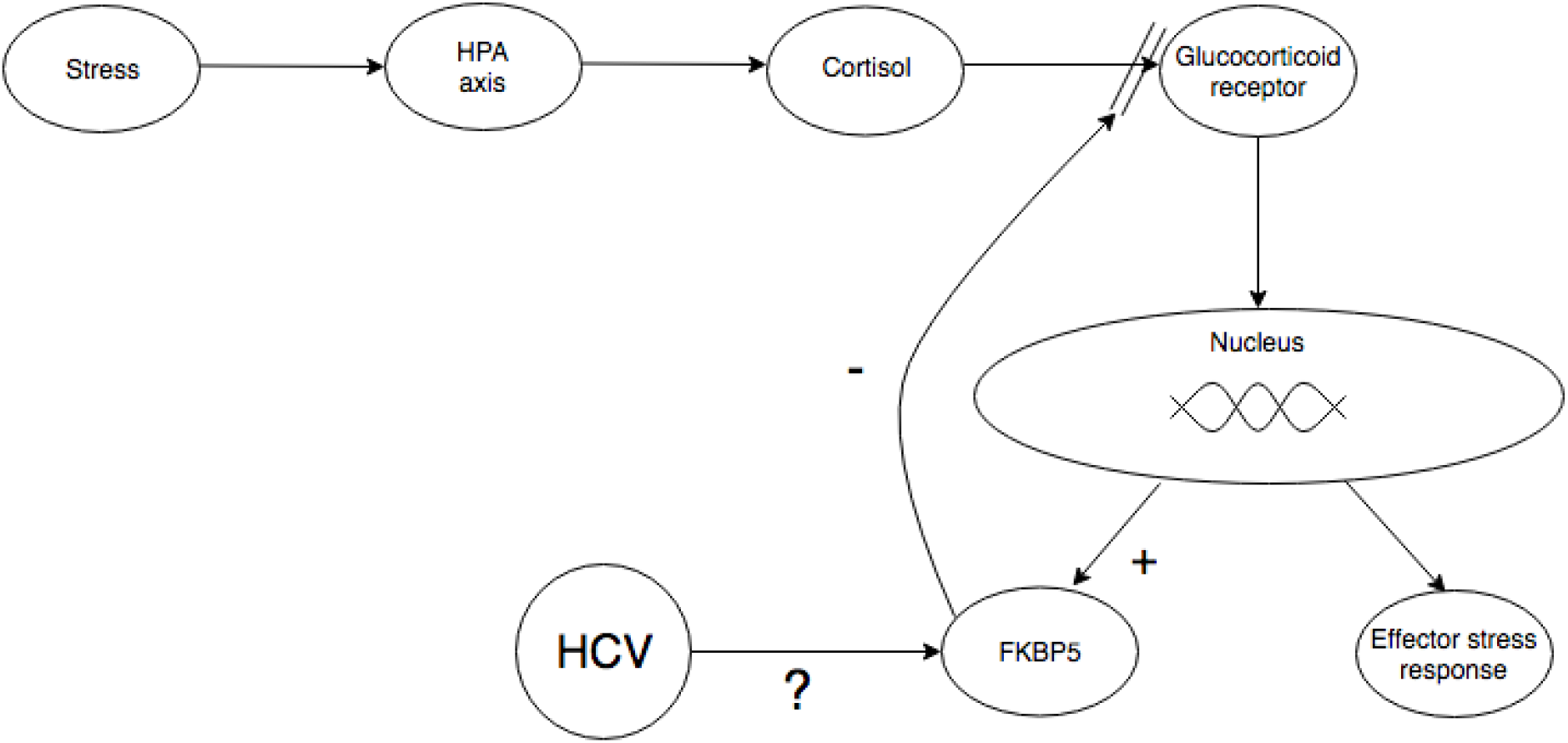
The relationship between FKBP5 and the glucocorticosteroid receptor. HPA; hypothalamic-pituitary-adrenal; HCV; hepatitis C virus.

The minor T allele of the rs1360780 SNP confers a high induction phenotype frequently associated with vulnerability to stress-induced mood and anxiety disorders. This association was first described in a case-cohort study in which TT homozygous subjects with depressive illness were identified as having more depressive episodes but also a better response to antidepressant treatment (irrespective of drug class)^10^. Furthermore, the authors found that those with TT rs1360780 genotype had twice the intracellular lymphocyte concentration of FKBP51 in spite of no corresponding increase in mRNA transcription (implying either enhanced translation or greater protein stability). Finally, they had higher GR resistance, as shown by a lower adrenocorticotropic hormone (ACTH) response to the combined dexamethasone-suppression/corticotrophin releasing hormone stimulation test commonly used to detect pseudo-Cushing states. Other groups have identified similar associations with unipolar depression; one prospective gene-environment study identified an interaction between TT allele, traumatic life events and the risk of first occurrence of a major depressive episode^14^. Similar associations have been seen in other stress related disorders including bipolar depression, post-traumatic stress disorder, suicidal ideation and psychosis.

Individuals chronically infected by the human immunodeficiency virus (HIV) have excess prevalence of depression and in a study of the cortical grey matter of infected patients, CC rs1360780 and CC rs3800373 FKBP5 genotypes were associated with major depression and depression/psychosis. There was elevated neuronal FKBP51 protein and transcript levels compared to non-depressed HIV controls^15^. Though it is unclear if there is a direct mechanism attributable to HIV in altering FKBP5 gene expression, it is postulated to be due to chronic systemic inflammation due to the virus^12^

No studies to date have examined the relationship between hepatitis C and FKBP5. This pilot study investigates the frequency of FKBP5 SNP polymorphisms, peripheral blood FKBP5 mRNA transcript levels and peripheral FKBP51 gene product levels in a cohort of CHC patients with depression compared to those without depression.

## METHODS

### Recruitment

Patients aged between 18 to 75 years with CHC (defined as detectable viral RNA for more than 6 months) were recruited prospectively from two tertiary referral centres. Exclusion criteria included decompensated cirrhosis (Child-Turcotte-Pugh class B or C), primary language other than English (as patients were required to complete multiple English language questionnaires), patients receiving interferon-based therapy, inability to provide voluntary written consent, inability to complete questionnaires and pregnancy. This study was approved by the Melbourne Health Office for Research Human Ethics committee and the Eastern Health Office of Research and Ethics. All subjects gave written informed consent prior to taking part in the study. All research was performed in accordance with relevant guidelines and regulations.

### Clinical and psychosocial data

Subjects completed surveys detailing highest level of education, personal and parents’ country of birth, medical comorbidities including hepatitis B and HIV co-infection, presence of heart disease, diabetes, arthritis and thyroid abnormalities. Current or past use of prescription drug and illicit substances including non-opioid analgesia, alcohol, amphetamines, benzodiazepines, cannabis, cocaine, ecstasy and opiates such as heroin was recorded.

### Psychiatric and Quality of Life Questionnaires

Psychiatric and Quality of Life morbidity were assessed using the following validated tools:

#### Hospital Anxiety and Depression Scale (depression subscore) – (HADS-D)

self-assessment scale designed to detect depression and anxiety disorders in non-psychiatric hospital settings^16^. The HADS depression subscore (HADS-D) correlates well with DSM-IV criteria for depression, and a cut off of 8 (out of 21) achieved optimal sensitivity and specificity of 0.8-0.9 for ‘possible’ cases of depression in a review^17^. We used a higher cut-off ≥11 to define ‘depressed’ subjects so as to improve specificity.

#### Modified Fatigue Impact Scale (MFIS)

21-item survey pertaining to the impact of fatigue on quality of life in the preceding four weeks with responses ranked on a Likert scale (0-4) according to frequency of symptoms (where “0” indicates “never” and “4” indicates “almost always”). The maximum score is 84 and lower scores denote lower impact of fatigue on daily activity. It has been validated in patients with chronic liver disease^18^.

#### Short Form-36 (SF-36) quality of life questionnaire scale

A multidimensional questionnaire comprising 36 items pertaining to health-related quality of life (HRQoL) spanning eight different “scales”: physical functioning, role-physical, bodily pain, general health, vitality, social function, role-emotional and emotional wellbeing^19^. Each scale is individually standardised to an Australian reference population and weighted so as to calculate aggregate Physical (PCS) and Mental (MCS) Component Summary measures^20,21^. Higher scores denote better HRQoL. Lower scores have been found in CHC patients compared to patients with chronic hepatitis B infection and is apparent irrespective of active drug use^22^. An improvement in SF-36 scores has been demonstrated after successful eradication of CHC^23^.

### Laboratory methods

Whole blood was collected for FKBP5 mRNA quantification, FKBP51 protein quantification. DNA was extracted using standard protocols. Six SNPs (rs1360780, rs1754246, rs3777747, rs3800373, rs737054 and rs9380525) within FKBP5 including the 5 kilobase flanking regions were selected using a r-squared threshold of 0.80 and minor allele frequency threshold of 15% based on the 1000 Genome Project’s European population^24^. Genotyping was performed using the iPLEX® Gold SNP genotyping kit (Agena, San Diego, USA) and the software and equipment provided with the MassARRAY® platform (Agena, San Diego, USA).

## Statistical analysis

Data were assessed for normality and presented at mean ± standard deviation (SD) for normally distributed quantitative variables, and median (interquartile range) for non-normally distributed variables while qualitative variables were presented as frequency (percentage). Baseline characteristics were compared using independent samples t test for parametric continuous variables and chi square test for categorical variables. Spearman rho correlation was used to assess correlation between questionnaire scores and FKBP51 protein levels. Chi square testing was used to compare genotype frequency between the groups and Kruskal-Wallis testing to determine associations between SNPs and mRNA and protein levels. All statistical analysis was performed using SPSS (Version 21.0) for Windows (SPSS Inc., Chicago, IL, USA).

## RESULTS

### Descriptive analyses

Fifty-three patients met the inclusion criteria and were invited to participate (figure 2). Forty-one patients were included in the final analysis after 12 were excluded because of incomplete questionnaires (n=7) or incomplete blood specimen collection (n=5). No relevant significant differences were seen when the 5 subjects with missing blood specimens were compared to the included 41 patients (supplementary tables 1-3). Comparison of descriptive data in the 7 with incomplete questionnaires could not be undertaken.

**Table 1:** Descriptive characteristics

|  |  | **Depressed group**<br>**n =13** | **Control Group**<br>**n= 28** | **p** |
| --- | --- | --- | --- | --- |
| **Age<sup>a</sup>** |  | 54.5 (13) | 53.6 (12.6) | 0.84 |
| **BMI<sup>a</sup>** |  | 25.4 (4.5) | 26 (4.3) | 0.65 |
| **Gender<sup>b</sup>** | **Male** | 9 (69) | 19 (68) | 0.93 |
|  | **Female** | 4 (31) | 9 (32) |  |
| **Highest level of education**<br>**achieved<sup>b</sup>** | **Less than year**<br>**12** | 9 (69.2) | 16 (57.1) | 0.51 |
|  | **Year 12 or**<br>**higher** | 4 (30.8) | 12 (42.9) |  |
| **Country of birth<sup>b</sup>** | **Australia** | 9 (69) | 20 (71) | 0.89 |
|  | **Overseas** | 4 (31) | 8 (29) |  |
| **HADS-D<sup>c</sup>** |  | 13 (3) | 4 (4) | **<0.01** |
| **HADS-A<sup>c</sup>** |  | 11 (8) | 5 (5) | **0.03** |
| **Modified Fatigue Impact Scale<sup>a,\*</sup>** |  | 57 (14) | 31 (16) | **<0.01** |
| **Short Form-36<sup>a, ^</sup>** | **Physical function** | 55 (27) | 74 (23) | **0.03** |
|  | **Role physical** | 35 (23) | 63 (25) | **<0.01** |
|  | **Bodily pain** | 51 (25) | 69 (25) | **0.04** |
|  | **General health** | 32 (11) | 49 (14) | **<0.01** |
|  | **Vitality** | 32 (17) | 52 (21) | **<0.01** |
|  | **Social function** | 38 (18) | 69 (25) | **<0.01** |
|  | **Role emotional** | 21 (30) | 69 (24) | **<0.01** |
|  | **Emotional**<br>**wellbeing** | 40 (16) | 70 (18) | **<0.01** |
|  | **Physical**<br>**Component**<br>**Summary** | 28.63 (12.52) | 38.18 (11.80) | **0.02** |
|  | **Mental Component**<br>**summary** | 30.07 (10.10) | 43.70 (11.21) | **<0.01** |

Data are expressed as <sup>a</sup>mean (standard deviation), <sup>b</sup>frequency (percentage), <sup>c</sup> median (IQR). \* a lower score denotes lower impact of fatigue on daily activity. ^ a higher score indicates better health-related quality of life. BMI = body mass index (kg/m<sup>2</sup>). HADS-A = Hospital Anxiety and Depression Scale anxiety subscore, HADS-D = Hospital Anxiety and Depression Scale depression subscore

**Fig. 2.**
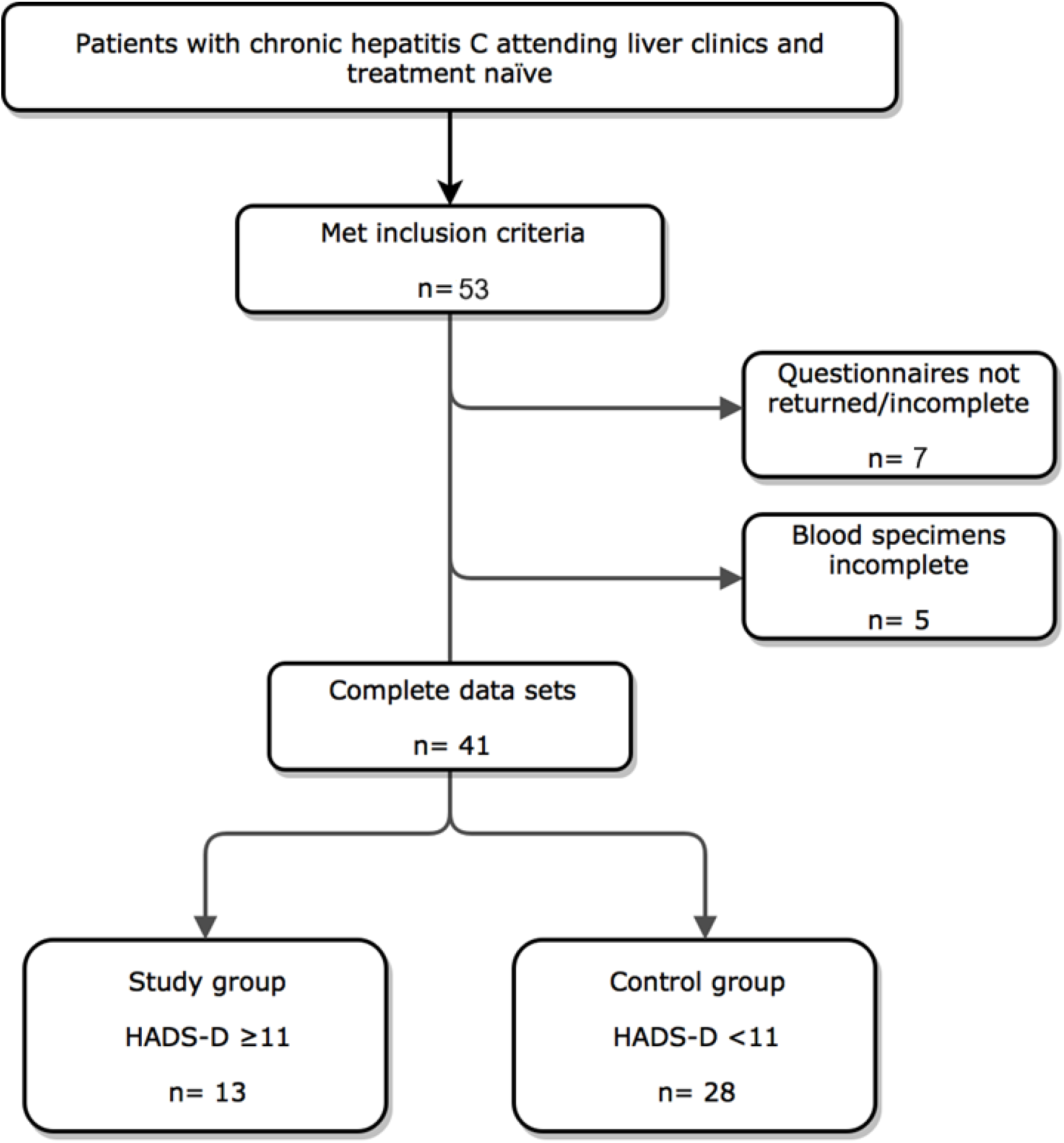
Study design. CLD chronic liver disease, HADS-D Hospital Anxiety and Depression Scale (depression subscore), MFIS; Modified Fatigue Impact Scale, SF36; Short Form 36 quality of life questionnaire scale.

The 41 with complete data were divided into two groups according to HADS-D sub score. Participants in the ‘depression’ group scored ≥ 11 (n= 13) while patients in the ‘control’ group had scores <11 (n = 28). The median HADS-D sub score was 13 in the ‘depression’ group and 4 in the control group. Patient characteristics are presented in Table 1. Sixty-eight percent were male (n = 28) and the mean age was 54 years. No significant differences between the depression and control groups were found in terms of age, body mass index, gender, highest level of education completed nor country of birth (Australia or overseas). HADS-D along with HADS-A sub scores were significantly different between the groups (p <0.01 and 0.03 respectively). MFIS scores were significantly lower in the non-depressed group (p <0.01 while SF-36 scores in all eight domains in addition to the aggregate physical and mental component summary scores, PCS and MCS were lower in the depressed group (table 1).

CHC genotypes 1 and 3 were the most prevalent with no patients having genotypes 2, 5 or 6; no significant differences in CHC genotype were seen between the two groups. Only the control group had individuals with hepatitis B co-infection but this did not reach statistical significance (p 0.54). Medical co-morbidities (supplementary table 2) did not differ between groups however those in the ‘depressed’ group were almost twice more likely to be diagnosed with depression in the past (p=0.018). There was also 2.6 times higher prevalence of past diagnosis of anxiety disorders in the depression group, compare to the controls but this did not reach statistical significance (p = 0.073). There were no differences in the rate of past or current substance use.

**Table 2.**
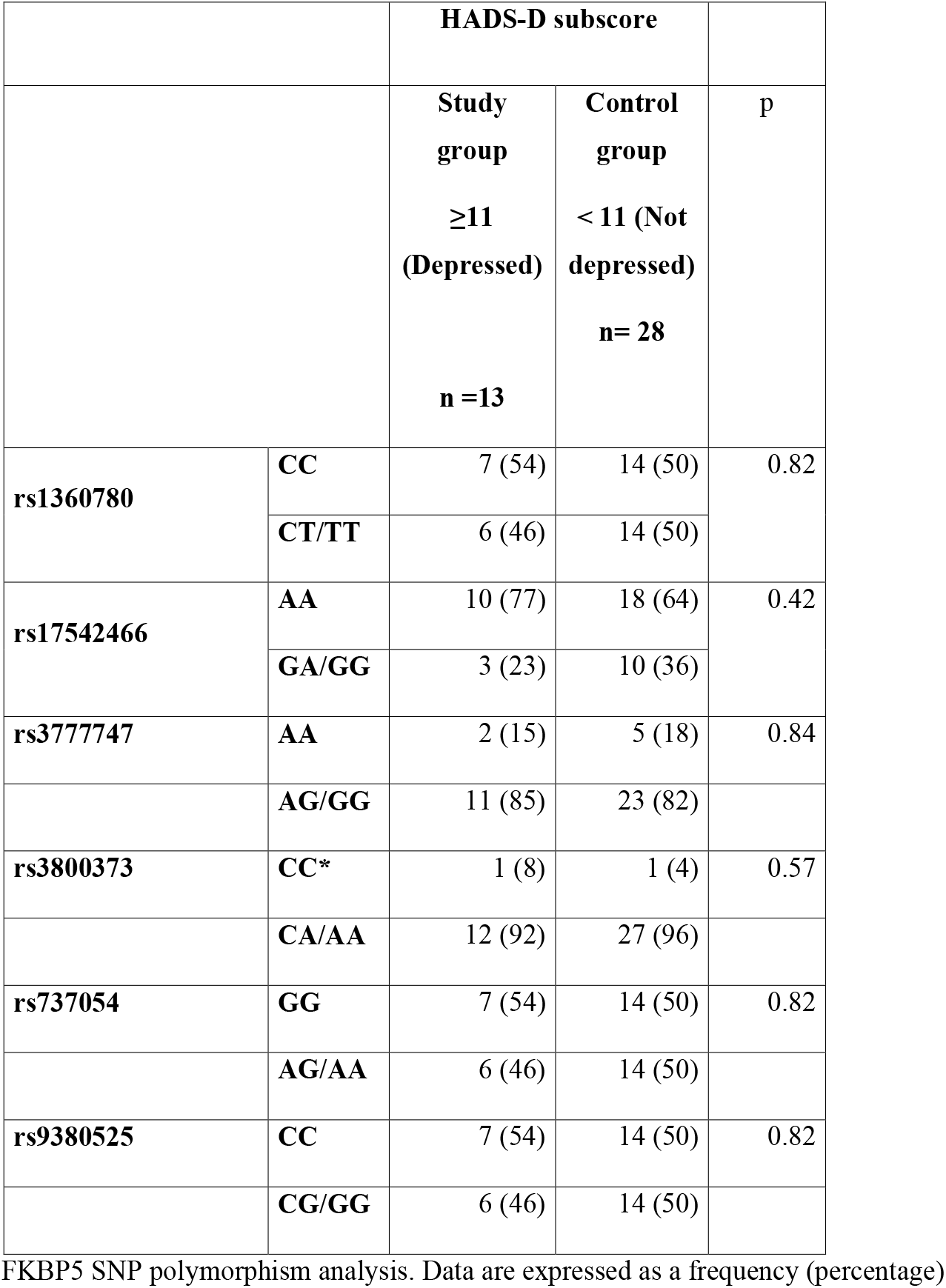
FKBP5 SNP polymorphism analysis

|  |  | **HADS-D subscore** |  |  |
| --- | --- | --- | --- | --- |
|  |  | **Study group**<br>**≥11 (Depressed)**<br>**n =13** | **Control group**<br>**< 11 (Not depressed)**<br>**n= 28** | **p** |
| **rs1360780** | **CC** | 7 (54) | 14 (50) | 0.82 |
|  | **CT/TT** | 6 (46) | 14 (50) |  |
| **rs17542466** | **AA** | 10 (77) | 18 (64) | 0.42 |
|  | **GA/GG** | 3 (23) | 10 (36) |  |
| **rs3777747** | **AA** | 2 (15) | 5 (18) | 0.84 |
|  | **AG/GG** | 11 (85) | 23 (82) |  |
| **rs3800373** | **CC\*** | 1 (8) | 1 (4) | 0.57 |
|  | **CA/AA** | 12 (92) | 27 (96) |  |
| **rs737054** | **GG** | 7 (54) | 14 (50) | 0.82 |
|  | **AG/AA** | 6 (46) | 14 (50) |  |
| **rs9380525** | **CC** | 7 (54) | 14 (50) | 0.82 |
|  | **CG/GG** | 6 (46) | 14 (50) |  |

FKBP5 SNP polymorphism analysis. Data are expressed as a frequency (percentage)

### Peripheral FKBP51 protein levels

Peripheral blood FKBP51 protein levels ranged between 0.211 to 3.385 ng/ml with a median of 1.132 (Q1-Q3: 1.037-1.496) ng/ml. The median FKBP51 level was significantly lower in the depression group 0.911 (0.806-1.072) ng/ml compared to 1.252 (1.10-1.743) ng/ml in the control group (p<0.01). A moderate negative correlation (rho= -0.53, p <0.01) was identified between HADS-D sub-score and FKBP51 levels (figure 3) as well as HADS-A sub-score and FKBP51 (rho= -0.31, p=0.04, figure 4). A weakly negative correlation (figure 5) was also seen between MFIS and FKBP51 (rho= -0.34, p =0.04). Of the individual SF36 domains, FKBP51 positively correlated with social functioning (r= 0.31, p = 0.04) and emotional wellbeing (rho= 0.39, p = 0.01).

**Fig. 3.**
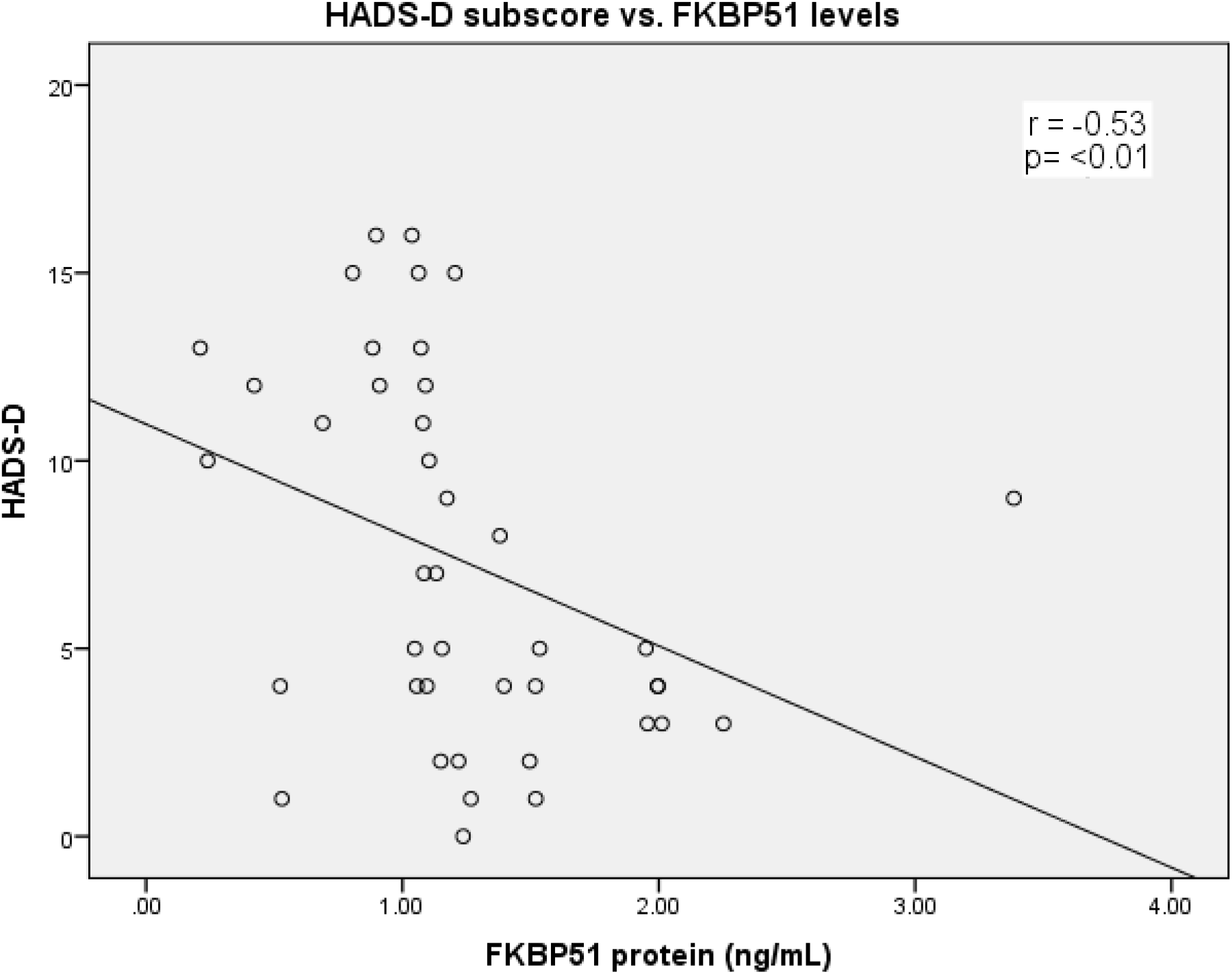
Correlation between HADS-D subscore and FKBP51 levels. HADS-D; Hospital Anxiety and Depression Scale (depression subscore)

**Fig. 4.**
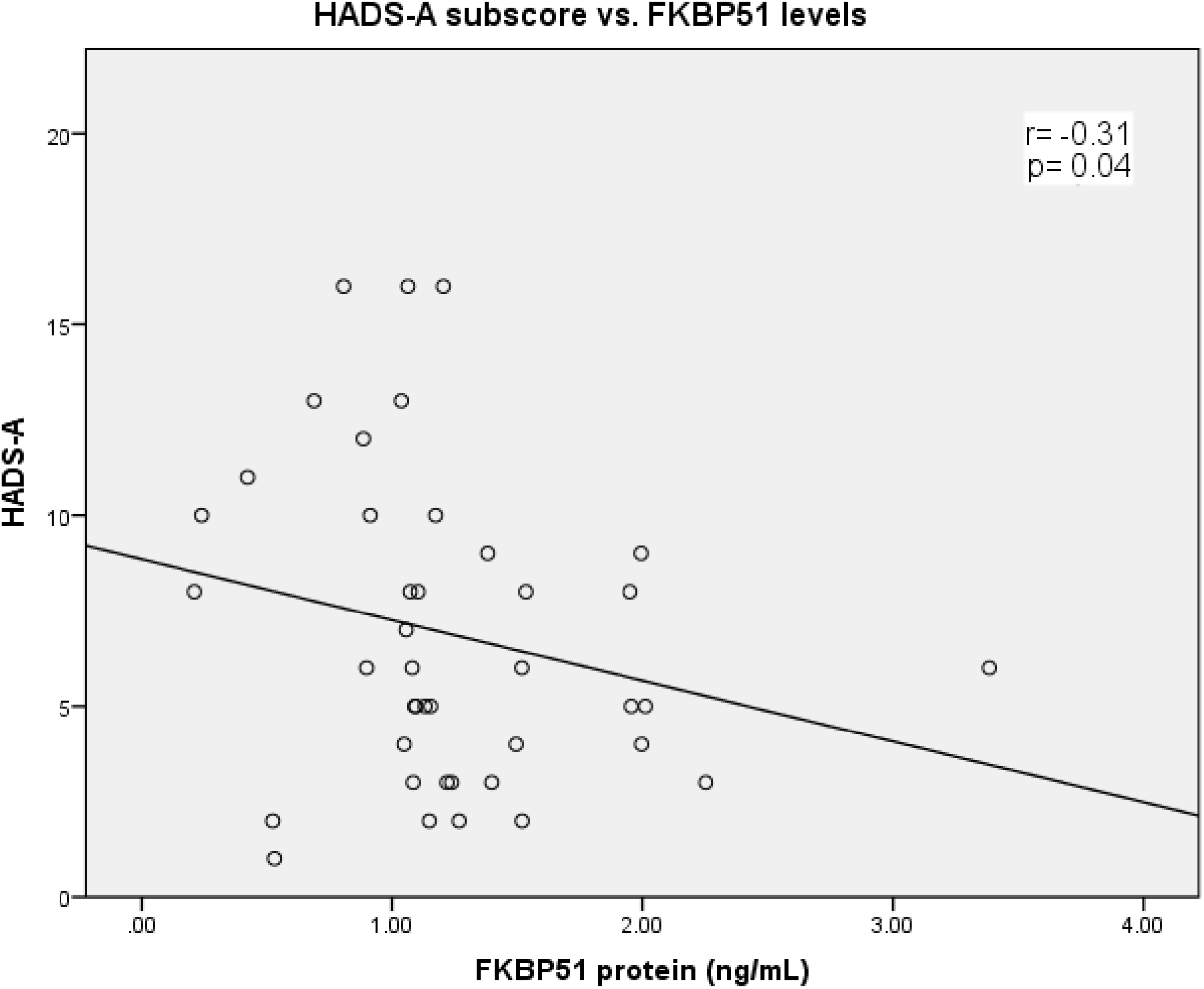
Correlation between HADS-A subscore and FKBP51 levels. HADS-A Hospital Anxiety and Depression (anxiety subscore)

**Figure 5:**
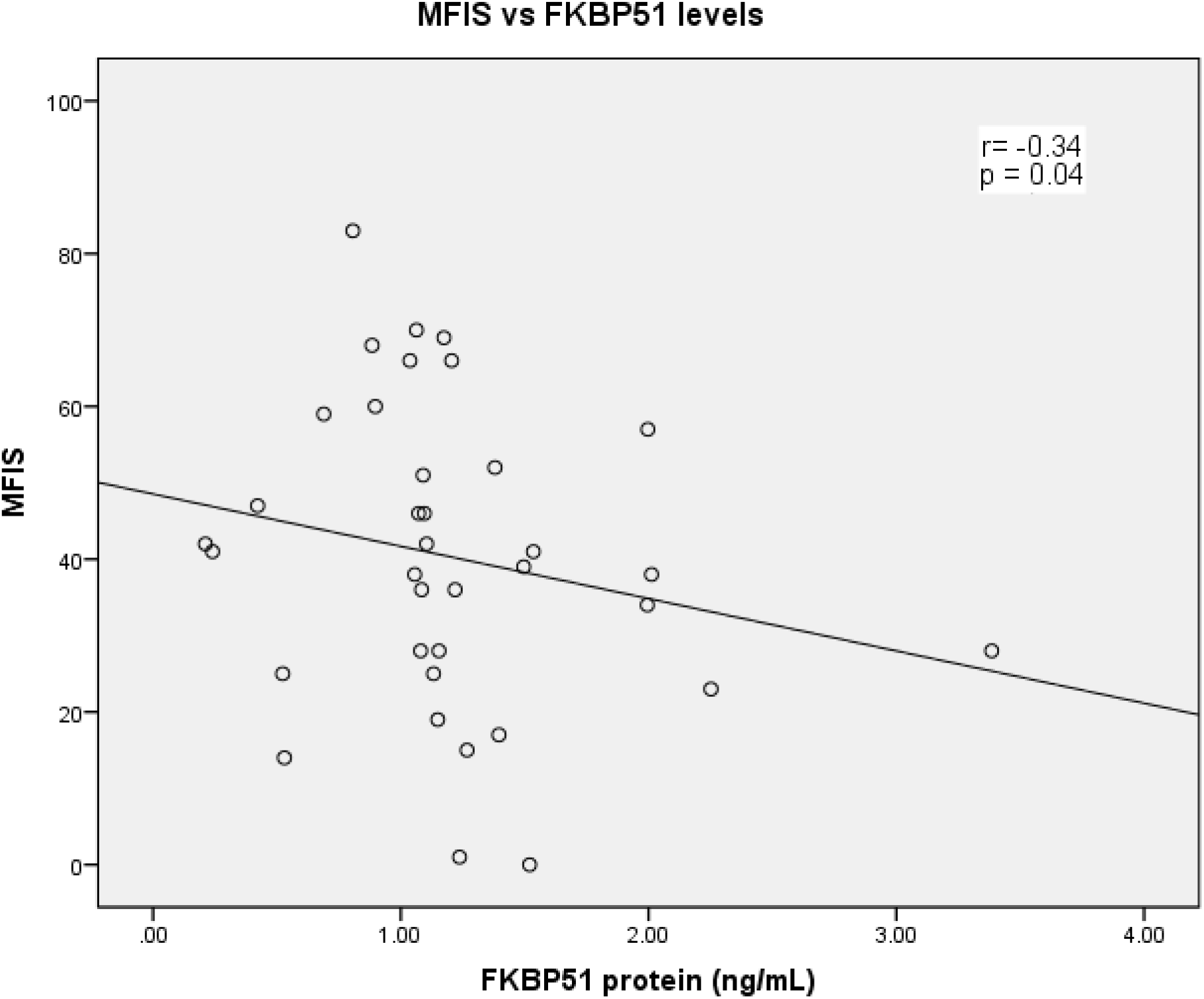
Correlation between MFIS and FKBP51 protein level. MFIS; Modified Fatigue Impact Scale.

### FKBP5 mRNA levels

FKBP5 mRNA PCR cycle thresholds were normalised to the reference gene HPRT1 and delta-delta CT levels calculated. The average expression fold change in FKBP5 mRNA in the depressed group was lower at 0.76 (with reference to HPRT1), compared to controls with a fold change of 1.40 (ratio of depression group to controls of 0.54). This did not reach statistical significance according to non-parametric Mann-Whitney U test, p=0.133.

### FKBP5 SNP polymorphism analysis

No significant differences in genotype frequency were seen between the depression and control groups (table 2). Kruskal-Wallis testing did not reveal any significant relationship between SNPs and FKBP5 mRNA levels nor FKBP51 protein levels. No associations were observed between FKBP5 and genetic data. (p>0.05 for all comparisons)

## DISCUSSION

This is the first study to examine the interaction between CHC and FKBP5. We hypothesised that high induction FKBP5 SNPs would be associated with the depressive phenotype in patients with CHC and that peripheral FKBP51 protein levels would be correspondingly elevated. However, no interaction between specific FKBP5 SNPs and HADS-D depression score was found in our study. We identified a significant, inverse correlation between the depression and anxiety sub-scores with peripheral blood levels of FKBP51 along with a trend towards lower mRNA levels. This was unexpected given other studies have demonstrated upregulation of this protein in accordance with HPA axis upregulation in the depressed state. The reasons for this are unclear but one reason for the discrepancy may have been due to how FKBP5 expression is measured, knowing that it may vary between different cell types and peripheral levels may not reflect cortical grey matter expression.

It may be expected that a direct correlation exists between liver disease activity and the severity of depressive illness, but the relationship is not straightforward. There is weak positive correlation between hepatic inflammation and non-significant correlation with hepatic fibrosis on biopsy with depressive symptoms^25-28^. The presence of hepatic encephalopathy can confound assessment of mood, along with the physical restrictions of large volume ascites and sarcopaenia impacting on quality of life and mood. In our study we controlled for this by excluding patients with decompensated liver disease.

Our study has limitations. As with all case-control gene association studies, case and control definitions may misclassify patients and result in biased results. To define our cases we utilised a HADS-D sub-score of ≥11; for logistical reasons, formal assessment by a qualified psychiatrist was not possible but would have improved case identification. Whilst the HADS is a well-validated tool, its original intention was for screening of patients for potential affective or anxiety-spectrum disorders in hospital settings rather than as a *diagnostic* tool. Thus subjects categorised as ‘depressed’ according to this score were not necessarily depressed by traditional clinic-pathologic criteria. We tried to minimise for this effect by raising the cut-off to 11 compared to the usual 8 applied in the clinical setting so as to improve specificity. Additionally, the retrospective nature of the psychosocial questionnaires means they may be subject to response bias.

Confounding in the form of physical comorbidities may also affect reported depressive symptoms. This is well-established in large cohort studies of subjects with CHC in which higher comorbidity scores were positively correlated with greater depressive symptoms^29^. In our study we did not find any significant difference in comorbidities between groups so confounding has been minimised.

We had a small number of patients whose data were analysed and there was an unexpectedly high rate of unusable data (arising from incomplete questionnaires and blood testing) which may have reduced our power to detect significant differences between the two groups. Spurious associations between SNPs and symptoms due to ethnic differences between cases and controls was minimised as there were similar number of patients born in Australia vs. overseas between the two groups. Though it has been noted in the literature that HPAA dysregulation associated with particular FKBP5 SNPs has been observed amongst a wide range of different ethnicities^30^.

Finally, we did not assess serum cortisol levels or administer dexamethasone/corticotropin releasing hormone (CRH) stimulation testing as it would have been useful to clarify if GR responsiveness was preserved (normal ACTH response to CRH) in light of low FKBP5 levels or if it remained insensitive (low ACTH response to CRH), thus confirming chronic hypercortisolaemia and suggesting that a mechanism (or several) separate to FKBP5 are impacted by CHC.

This study has explored the relationship between hepatitis C virus and a candidate gene for depression. With the increasing availability of direct-acting antivirals and real possibility of eradicating the virus within the next 30 years, barriers to treatment such as psychiatric illness can preserve reservoirs of virus in socially isolated patients. Therefore, greater understanding of the pathogenic role the virus plays in the pathogenesis of depression will remain an important goal of future research.

## Data Availability

Data is available from the authors.
There is no external database

## Abbreviations

ACTH: Adrenocorticotropic hormone BMI Body mass index
CHC: Chronic hepatitis C
CRH: corticotropin releasing hormone
DNA: Deoxyribonucleic acid
GR: Glucocorticosteroid receptor
HADS-A: Hospital Anxiety and Depression Scale anxiety sub-score
HADS-D: Hospital Anxiety and Depression Scale depression sub-score
HRQoL: Health-related quality of life
HIV: Human Immunodeficiency Virus
HPAA: Hypothalamic-pituitary-adrenal axis
MCS: Mental Component Summary
mRNA: Messenger ribonucleic acid
MFIS: Modified Fatigue Impact Scale
PCS: Physical Component Summary
SD: Standard deviation
SF-36: Short Form-36 quality of life questionnaire scale
SNP: Single nucleotide polymorphisms

## REFERENCES

1. Stewart B, Mikocka-Walus A, Morgan J, et al. Anxiety and depression in Australian chronic hepatitis C outpatients: prevalence and predictors. Australas Psychiatry. 2012;20(6):496–500.

2. Sood S, Wong D, Holmes A, Everall I, Saling M, Nicoll A. Depression in a Real World Population of Hepatitis C Patients. 2014;1(2):1–3.

3. Yeoh SW,, Holmes ACN, Saling MM, Everall IP, Nicoll AJ. Depression, fatigue and neurocognitive deficits in chronic hepatitis C. Hepatol Int LID - 10.1007/s12072-018-9879-5 [doi]. (1936-0541 (Electronic)).

4. Rogal SS, McCarthy R, Reid A, et al. Primary Care and Hepatology Provider-Perceived Barriers to and Facilitators of Hepatitis C Treatment Candidacy and Adherence. Dig Dis Sci. 2017;62(8):1933–1943.

5. Laskus T, Radkowski M, Bednarska A, et al. Detection and analysis of hepatitis C virus sequences in cerebrospinal fluid. J Virol. 2002;76(19):10064–10068.

6. Radkowski M, Wilkinson J, Nowicki M, et al. Search for hepatitis C virus negative-strand RNA sequences and analysis of viral sequences in the central nervous system: evidence of replication. J Virol. 2002;76(2):600–608.

7. Fletcher NF, Wilson GK, Murray J, et al. Hepatitis C virus infects the endothelial cells of the blood-brain barrier. Gastroenterology. 2012;142(3):634–643 e636.

8. Forton DM, Hamilton G, Allsop JM, et al. Cerebral immune activation in chronic hepatitis C infection: a magnetic resonance spectroscopy study. J Hepatol. 2008;49(3):316–322.

9. Byrnes V, Miller A, Lowry D, et al. Effects of anti-viral therapy and HCV clearance on cerebral metabolism and cognition. Journal of hepatology. 2012;56(3):549–556.

10. Binder EB, Salyakina D, Lichtner P, et al. Polymorphisms in FKBP5 are associated with increased recurrence of depressive episodes and rapid response to antidepressant treatment. Nat Genet. 2004;36(12):1319–1325.

11. O’Leary JC, 3rd, Zhang B, Koren J, 3rd, Blair L, Dickey CA. The role of FKBP5 in mood disorders: action of FKBP5 on steroid hormone receptors leads to questions about its evolutionary importance. CNS Neurol Disord Drug Targets. 2013;12(8):1157–1162.

12. Tatro ET, Nguyen TB, Bousman CA, et al. Correlation of major depressive disorder symptoms with FKBP5 but not FKBP4 expression in human immunodeficiency virus-infected individuals. J Neurovirol. 2010;16(5):399–404.

13. Lekman M, Laje G, Charney D, et al. The FKBP5-gene in depression and treatment response--an association study in the Sequenced Treatment Alternatives to Relieve Depression (STAR*D) Cohort. Biol Psychiatry. 2008;63(12):1103–1110.

14. Zimmermann P, Bruckl T, Nocon A, et al. Interaction of FKBP5 gene variants and adverse life events in predicting depression onset: results from a 10-year prospective community study. Am J Psychiatry. 2011;168(10):1107–1116.

15. Tatro ET, Everall IP, Masliah E, et al. Differential expression of immunophilins FKBP51 and FKBP52 in the frontal cortex of HIV-infected patients with major depressive disorder. J Neuroimmune Pharmacol. 2009;4(2):218–226.

16. Zigmond AS, Snaith RP. The hospital anxiety and depression scale. Acta Psychiatr Scand. 1983;67(6):361–370.

17. Bjelland I, Dahl AA, Haug TT, Neckelmann D. The validity of the Hospital Anxiety and Depression Scale. An updated literature review. J Psychosom Res. 2002;52(2):69–77.

18. Lundgren-Nilsson A, Tennant A, Jakobsson S, Simren M, Taft C, Dencker A. Validation of Fatigue Impact Scale with various item sets - a Rasch analysis. Disabil Rehabil. 2017:1–7.

19. Measuring Functioning and well-being: The medical outcomes study approach; Anita L. Stewart and John E. Ware, Jr (editors). Duke university press, Durham and London, 1992. No. of pages: 449. ISBN 0–8223–1212–3. Price: US$55. Psycho-Oncology. 1995;4(2):163–163.

20. Australian Bureau of Statistics. 1995 National Health Survey – SF-36 Population Norms. In. Vol ABS Catalogue No. 4399.0. Australia: ABS; 1997.

21. Ware JE, New England Medical Center H, Health I. SF-36 physical and mental health summary scales : a user’s manual. Boston: Health Institute, New England Medical Center; 1994.

22. Foster GR, Goldin RD, Thomas HC. Chronic hepatitis C virus infection causes a significant reduction in quality of life in the absence of cirrhosis. Hepatology (Baltimore, Md). 1998;27(1):209–212.

23. Bonkovsky HL, Woolley JM. Reduction of health-related quality of life in chronic hepatitis C and improvement with interferon therapy. The Consensus Interferon Study Group. Hepatology (Baltimore, Md). 1999;29(1):264–270.

24. The Genomes Project C. A global reference for human genetic variation. Nature. 2015;526:68.

25. Zelber-Sagi S, Toker S Fau - Armon G, Armon G Fau - Melamed S, et al. Elevated alanine aminotransferase independently predicts new onset of depression in employees undergoing health screening examinations. (1469-8978 (Electronic)).

26. Ko FY, Yang Ac Fau - Tsai S-J, Tsai Sj Fau - Zhou Y, Zhou Y Fau - Xu L-M, Xu LM. Physiologic and laboratory correlates of depression, anxiety, and poor sleep in liver cirrhosis. (1471-230X (Electronic)).

27. Tomeno W, Kawashima K, Yoneda M, et al. Non-alcoholic fatty liver disease comorbid with major depressive disorder: The pathological features and poor therapeutic efficacy. J Gastroenterol Hepatol. 2015;30(6):1009–1014.

28. Youssef NA, Abdelmalek Mf Fau - Binks M, Binks M Fau - Guy CD, et al. Associations of depression, anxiety and antidepressants with histological severity of nonalcoholic fatty liver disease. (1478-3231 (Electronic)).

29. Boscarino JA, Lu M, Moorman AC, et al. Predictors of poor mental and physical health status among patients with chronic hepatitis C infection: the Chronic Hepatitis Cohort Study (CHeCS). Hepatology. 2015;61(3):802–811.

30. Zannas AS, Wiechmann T, Gassen NC, Binder EB. Gene-Stress-Epigenetic Regulation of FKBP5: Clinical and Translational Implications. (1740-634X (Electronic)).

